# Diagnosing COVID-19 in human serum using Raman spectroscopy: a preliminary study

**DOI:** 10.1101/2021.08.09.21261798

**Authors:** Ana Cristina Castro Goulart, Landulfo Silveira, Henrique Cunha Carvalho, Cristiane Bissoli Dorta, Marcos Tadeu T. Pacheco, Renato Amaro Zângaro

## Abstract

This preliminary study proposed the diagnosis of COVID-19 by means of Raman spectroscopy. Samples of blood serum from 10 patients positive and 10 patients negative for COVID-19 by RT-PCR RNA and ELISA tests were analyzed. Raman spectra were obtained with a dispersive Raman spectrometer (830 nm, 350 mW) in triplicate, being submitted to exploratory analysis with principal component analysis (PCA) to identify the spectral differences and discriminant analysis with PCA (PCA-DA) and partial least squares (PLS-DA) for classification of the blood serum spectra into Control and COVID-19. The spectra of both groups positive and negative for COVID-19 showed peaks referred to the basal constitution of the serum (mainly albumin). The difference spectra showed decrease in the peaks referred to proteins and amino acids for the group positive. PCA variables showed more detailed spectral differences related to the biochemical alterations due to the COVID-19 such as increase in lipids, nitrogen compounds (urea and amines/amides) and nucleic acids, and decrease of proteins and amino acids (tryptophan) in the COVID-19 group. The discriminant analysis applied to the principal component loadings (PC 2, PC 4, PC 5 and PC 6) could classify spectra with 87% sensitivity and 100% specificity compared to 95% sensitivity and 100% specificity indicated in the RT-PCR kit leaflet, demonstrating the possibilities of a rapid, label-free and costless technique for diagnosing COVID-19 infection.

## 1. Introduction

The current COVID-19 pandemic, the disease caused by the new coronavirus SARS-CoV-2, has caused enormous loss of lives [1] and negative economical effects, with significant increase in costs of medical inputs and equipments for the public and private health sectors worldwide caused by the high demand [2]. In addition, in the countries where the virus circulates, local governments oriented by World Health Organization (WHO) implemented spreading containment measures (closing borders and airports, closing trades and non-essential services, lockdown, social distance and massive testing) as well as the obligatory use of facial masks in public space [3, 4]. According to the WHO, a measure that has an important effect in controlling the transmission and consequently virus dissemination is the massive testing.

Vertical isolation or mitigation was initially adopted in the United Kingdom and the Netherlands, which later fell due to the large number of cases. Italy and Spain took a long time to determine horizontal social isolation, also called suppression, and account for the largest number of deaths. Suppression has been successfully adopted in China and South Korea [5]. In Brazil, a mixture of mitigation, suppression and lockdown has been adopted by most of the state governors and city mayors [6], while country’s president was not able to address the pandemic in satisfactorily way due to ideological positioning [7].

Public infectious disease control agents are faced with the need for a fast and reliable diagnostic technique for carriers of the SARS-CoV-2 virus in order to better manage the isolation of infected patients during the COVID-19 pandemic. Early diagnosis of COVID-19 also contributes in reducing the chances of sustained transmission of the disease in a condition of flattening of the contagion curve, in which the time to reach the “peak” and the herd immunity becomes longer [8, 9]. Another important fact is that in many countries like Brazil, testing is done only on symptomatic patients, leaving out a legion of asymptomatic individuals without diagnosis but with the virus [9, 10]. Also, there is a consensus that SARS-CoV-2 virus may become endemic with reinfections [11], and the COVID-19 disease may become a part of the “seasonal flu”.

In March 2020, WHO requested mass testing of the population in order to identify and isolate as many infected people as possible. One of the best examples of this testing came from South Korea, which did not quarantine, but tested millions of people who, together with others measures reduced the number of cases and deaths. Currently, Brazil performs 19.54 tests for every thousand inhabitants, behind countries like Chile 110.57/thousand inhabitants, Argentina 19.94/thousand inhabitants, Italy 76.10/ thousand inhabitants, USA 226.07/thousand inhabitants and United Kingdom 183.35/ thousand inhabitants [9, 10]. Therefore, the rapid and accurate diagnosis of COVID-19 is extremely important for the management of positive patients with isolation and adaptation of available hospital beds to the demand [12, 13].

The gold standard technique for the detection of the COVID-19 is based on the identification of the virus genetic material (ribonucleic acid – RNA) by RT-PCR (reverse transcription – polymerase chain reaction) technique in nasopharyngeal and oropharyngeal samples collected using a swab [14, 15]. Bronchoalveolar and sputum lavage materials may also be used in hospitalized patients [16]. However, the PCR technique has some disadvantages such as the need for specific physical space, equipments and trained personnel to perform the assay, higher costs compared to traditional serological tests for antibody detection, longer time to release the results (around 7 days), and they can present a false negative if the collection is performed improperly [15, 17].

Rapid and accurate diagnoses are essential to reduce the impact caused by tests with false negative results, requesting the development of new diagnostic tests [15, 18]. Spectroscopic methods for the analysis of biological materials offer several advantages over molecular biochemical methods, including speed in obtaining diagnostic information, no need for sample preparation or for use of reagents (label-free) [19]. Raman spectroscopy is a non-destructive analytical technique that provides information about the molecular composition of the materials studied, using a minimum amount of sample without the need for preparation, in addition to quick diagnosis that can be done in a few minutes using statistical and computational tools [20]. In fact, Raman spectroscopy has been used to quantify biomarkers of kidney disease such as urea and creatinine in serum [21], to quantify glucose and lipid components of sera [22] and to quantify prostate-specific antigen (PSA) values in sera of prostate cancer patients [23] aiming diagnosis.

Up to date, some authors have proposed the use of Raman spectroscopy for diagnosing viruses or virus diseases, particularly COVID-19. Desai et al. (2020) showed a preliminary report on the development of a statistical model to detect RNA viruses in saliva infected with viral particles by means of Raman spectroscopy, discussing possible applications and implications in the diagnosis of COVID-19 [24]. Masterson et al. (2020) proposed a method of “liquid biopsy” based on the evaluation of microRNA of cancers in plasma through surface-enhanced Raman spectroscopy (SERS) and plasmon-enhanced fluorescence (PEF), a method that could be expanded to the diagnosis of COVID-19 [25]. Also, Dou et al. (2020) showed that viruses could be identified in a label-free way by tip-enhanced Raman spectroscopy (TERS), by revealing protein secondary structure and amino acid composition of the virus surface [26]. Carvalho and Nogueira (2020) suggested in a “letter to the editor” that the vibrational techniques such as Raman and Fourier-transform infrared (FT-IR) spectroscopies could address the challenges for early and fast diagnosis of COVID-19 [27]. Khan and Rehman (2020) discussed the possibilities and challenges of developing a spectroscopic test methodology based on FT-IR and Raman spectroscopies to analyze COVID-19 from samples of serum, blood, saliva, urine and compare spectral results with the current PCR method [28].

The objective of this study is to show the preliminary results of a proposed methodology based on Raman spectroscopy of the serum and discriminant analysis to the diagnosis of serum samples of individuals with PCR positive for COVID-19 versus individuals with PCR negative. Also, we present an exploratory analysis based on principal component analysis (PCA) as a tentative biochemical description of the spectral differences of the serum positive for COVID-19 with regard the composition of proteins, amino acids, nucleic acids, lipids and carotenoids.

## 2. Materials and Methods

### 2.1 Serum samples

The study was approved by the Ethics and Research Committee of Universidade Anhembi Morumbi, under protocol No. 26691419.6.0000.5492.

Among several patients, 20 individuals were selected, 10 of them presenting positive results for COVID-19 (COVID-19 group) and the other 10 individuals presenting negative results for COVID-19 (Control group). Confirmation of these results was obtained by subjecting both groups to RT-PCR and ELISA (enzyme-linked immunonosorbent assay) tests. From each individual it was collected a sample of nasopharynx secretion, a sample of oropharynx, and 5 mL of blood at the CIPAX Diagnostic Medicine (São José dos Campos, SP, Brazil) [12, 15-17, 29].

The samples of secretion and blood were collected between the third and tenth day after the onset of the reported symptoms for the positive patients: fever (50%), cough (50%), sore throat (40%), anosmia (30%), malaise (30%), runny nose (30%), headache (20%), lack of appetite (10%), abdominal pain (10%), body pain (10%) and pneumonia (10%). To perform RT-PCR the material from the nasopharynx and oropharynx were collected by means of two polyester swabs and immediately placed in a tube with screw cap containing 3 mL of sterile saline solution (swabs and tube by Kolplast, São Paulo, SP, Brazil). The tubes were transported in an upright position at temperature of 2 °C to 8 °C until the sample was processed [16, 29, 30]. Whole blood was collected by means of venipuncture in a closed system using tubes with a separator gel (Vacuette^®^ Z Serum Sep Clot Activator, Greiner Bio-One, Americana, SP, Brazil). The blood tubes were subjected to centrifugation for 10 minutes at 3,000 RPM (model Elektra, Laborline, São Paulo, SP, Brazil) for serum extraction. Serum samples were used for serological tests [31].

The materials from nasopharynx and oropharynx were subjected to analyses performed using a RT-PCR assay (Abbott Real-Time SARS-CoV-2 kit, Abbott Laboratories, São Paulo, SP, Brazil), which characterizes the individuals as “detectable” and “undetectable” for COVID-19 disease [29]. The RT-PCR technique is considered the gold standard for the detection of COVID-19 for evidencing the RNA of the virus through the amplification of nucleic acid, with 95% sensitivity and 100% specificity as indicated in the leaflet for virus copies > 100 copies/mL [29]. The serum samples were submitted to the ELISA assay (Euroimmun Anti SARS-CoV-2 ELISA kit and Euroimmun Analyzer I, Euroimmun Brasil, São Caetano do Sul, SP, Brazil), for detection of IgA immunoglobulin. The results of ELISA tests were compared with RT-PCR and tests were in agreement for all patients. After the ELISA tests, the 20 serum samples were packed in thermal boxes, kept at 2 °C to 8 ºC and then submitted to Raman spectroscopy.

### 2.2 Raman spectroscopy

At the time of spectroscopic analysis, serum samples were passively thawed and 80 μL of serum was pipetted into an aluminum sample holder for reading the Raman spectra in triplicate. The spectra were obtained in a dispersive Raman spectrometer (model Dimension P1, Lambda Solutions Inc, MA, USA) with 830 nm excitation and 350 mW laser power. The spectrometer has a spectral resolution of 4 cm^-1^ in the spectral range between 400 and 1800 cm^-1^. The collection time for each spectrum was 30 s (3 s and 10 accumulations).

Before data analysis, Raman spectra were subjected to pre-processing to extract the fluorescence background (baseline) using a 7^th^ order polynomial. Other interferences such as cosmic rays were removed manually and the spectra were then normalized by the area under the curve (1-norm). Each triplicate spectrum was considered an individual sample in the data analysis.

### 2.3 Principal component analysis and classification

The Raman spectra were submitted to PCA in order to unveil the spectral differences between the two groups related to the changes in the biochemical constitution of the serum due to COVID-19. Also, the PCA variables (markedly the PCA loadings – PCs) were used in a discriminant analysis (DA) model. The DA model was also implemented via partial least squares (PLS) regression using the whole spectral information instead of the selected PCs [32]. Multivariate models such as PCA and PLS have been used for classification of Raman spectra of sera in normal and anemia [33] and for quantification of blood analytes in human serum for diagnosis [22, 23].

A Kolmogorov Smirnov normality test was applied to check the normality of the PCs and Student’s *t*-test (with Welch correction whenever needed) or Mann-Whitney U-test were applied to the PCs in order to identify significant differences between Control and COVID-19 groups. The *p*-value was considered significant when *p* < 0.05.

## 3. Results and Discussion

### 3.1 Raman spectra of serum samples of Control and COVID-19

Figure 1 presents the mean Raman spectra of the Control group and COVID 19 group, showing the difference spectra between them. Both groups showed spectral features referred to human serum, with peaks at 852, 941, 1003, 1319, 1341, 1450 and 1658 cm^-1^ assigned to albumin [34-37]. The difference spectra (scale multiplied by 3) showed positive and negative peaks along the spectral range; this suggest that the composition (proteins, lipids, amino acids) of both samples differ in some extent. For instance, the peaks at 621, 641, 939, 1003 and 1453 cm^-1^ can be assigned to proteins/amino acids; the peak at 1525 cm^-1^ can be assigned to carotenoids. The following exploratory analysis by PCA has been used to understand the nature of this biochemical difference.

**Figure 1.**
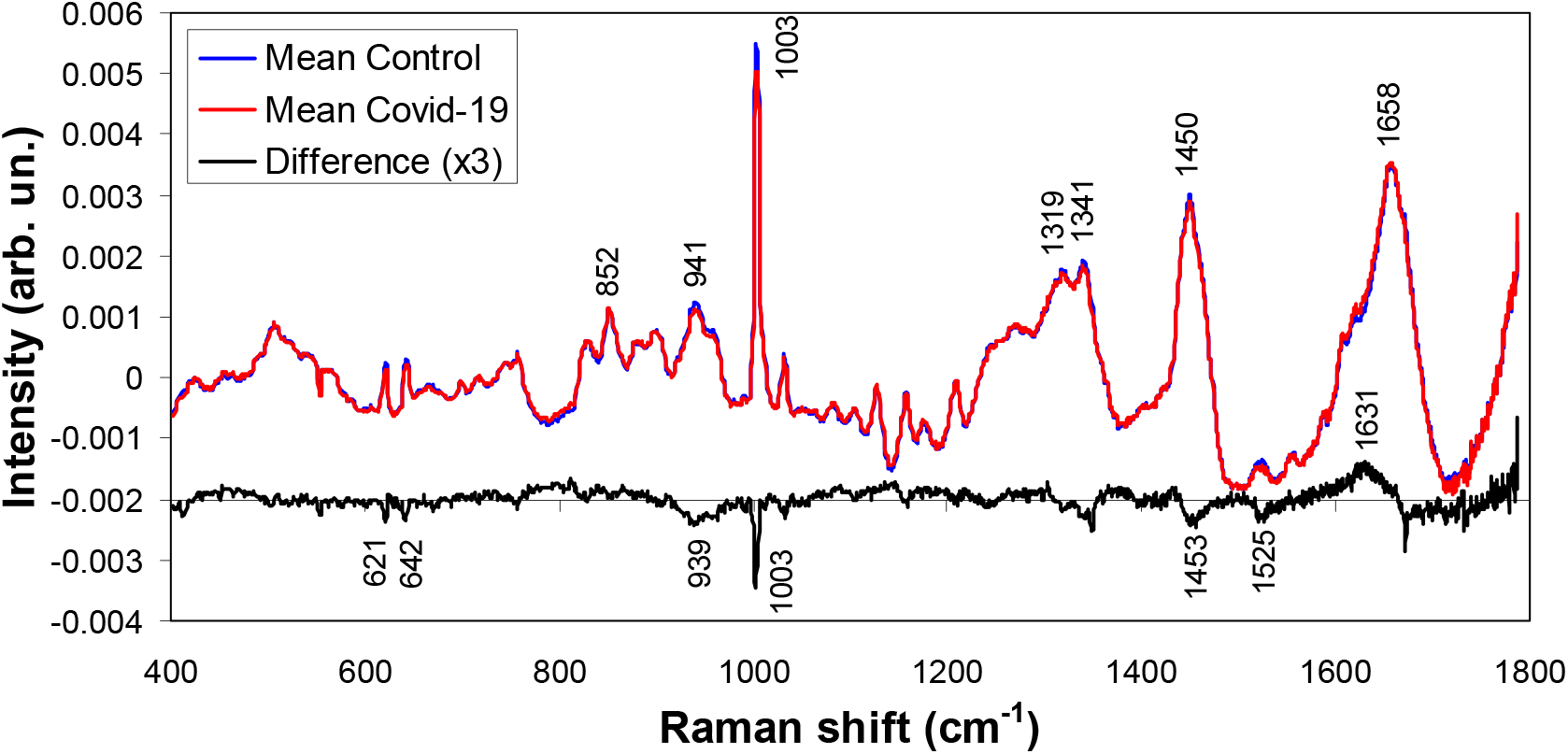
Mean Raman spectra of the blood serum of Control group and blood serum of patients with COVID-19 diagnosed by RT-PCR. Difference spectra (COVID-19 – Control) is also shown, with positive and negative peaks assigned to proteins, lipids, amino acids and nucleic acids as discussed in the text.

### 3.2 Exploratory analysis

The exploratory analysis used the PCA variables (Scores – spectral variances presented in the spectral data and PCs – intensities of each score in the original data) and Figure 2 shows the first 6 variables. These 6 Scores accounted for 99.4% of all spectral variation. Statistical significance of each PC in the group COVID-19 versus Control is also shown in Figure 2. The Score 1 represented the spectral features of serum (mainly albumin and γ globulin) [34-37], and both groups present similar constitution seen by the same intensity of PC 1 (not significant difference). On the other hand, Score 2 presented negative spectral features assigned to proteins (peaks at 939, 1003, 1454 and 1672 cm^-1^) [37-39] and carotenoids (peaks at 1159 and 1529 cm^-1^) [38], higher (negative) for the Control group, and positive spectral features which could be assigned to amines/amides and nucleic acids (peaks at 794, 1142, 1366 and 1630 cm^-1^) [38], higher for the COVID-19 group, as seen by PC 2 (statistically significant difference, Welch-corrected *t*-test, *p* < 0.0001). Most of these spectral features are the same as the ones found in the difference spectrum in Figure 1. A study showed that SARS-CoV2 affects the 40S ribosomal subunit by preventing its binding with messenger RNA and decreasing host protein production [40]. COVID-19 increases chemical mediators of inflammation derived from granules that are inside the cells such as tissue macrophages, endothelial cells, and leukocytes [41-43]. These chemical mediators include vasoactive amines (histamine), serotonin, arachidonic acid, platelet-activating factor, cytokines, neuropeptins. The increase in amine seen in the Raman spectra of positive patients can also be explained by the increase in transaminases (TGO and TGP), which are enzymes responsible for transferring an amine group from one amino acid to a hydrocarbon to form a different amino acid and by increasing urea (the main functional compound of amides) and nitrogenous amine creatine synthesized in the liver, kidneys and pancreas from amino acids glycine and arginine, also having methionine as a methyl donor [42, 44, 45]. Therefore these amines could be responsible for the Raman features referred to amines/amides which were present in the sera of the positive COVID-19 group. Also, the presence of features assigned to nucleic acids could be due to the presence of SARS-CoV-2 RNA in sera of positive patients.

**Figure 2.**
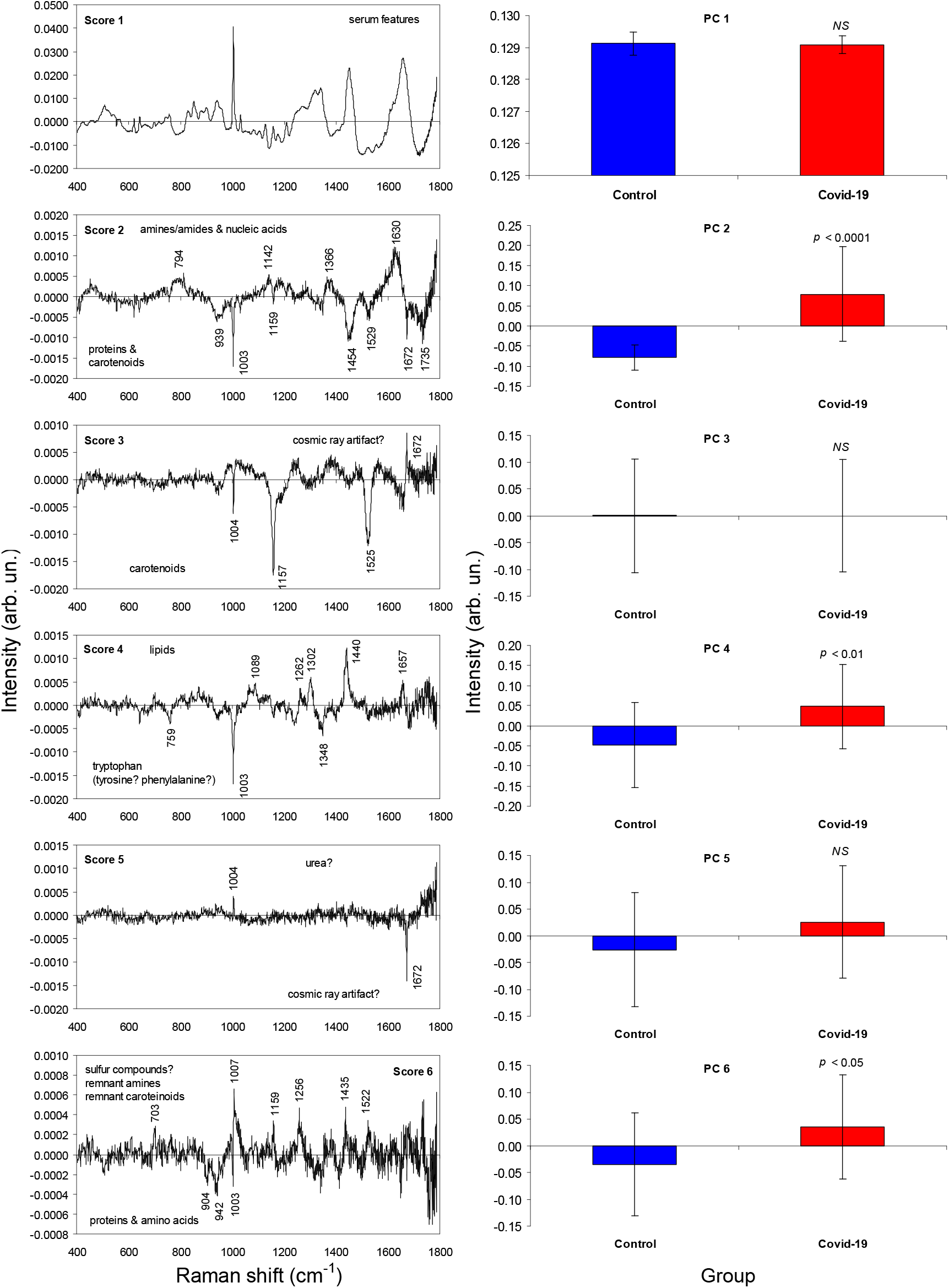
Plot of the first 6 principal component variables Scores and PCs. The features in Scores are assigned to serum constituents such as albumin, proteins, lipids, amino acids, nucleic acids, sulfur compounds and carotenoids. The intensities in PCs showed significant differences between groups for PC2, PC4 and PC6 (*t*-test and U-test, *p* < 0.05). *NS* = not significant.

Score 3 showed negative spectral features which are assigned to carotenoids (peaks at 1004, 1156 and 1525 cm^-1^) [38], but without difference between groups, as seen by PC 3 (not significant difference Mann-Whitney U-test, *p* = 0.387). Score 4 presented negative features assigned to amino acids – tryptophan (peaks at 759, 1003 and 1348 cm^-1^) [38], higher (negative) for the Control group, and positive features assigned to lipids (1089, 1262, 1302, 1440 and 1657 cm^-1^) [37-39], higher for COVID-19, as seen by PC 4 (statistically significant difference, Welch-corrected *t*-test, *p* < 0.01). Score 5 presented spectral features which may be assigned to urea [46] and possible cosmic ray artifact; the PC 5 showed no significant difference between groups (*t*-test, *p* = 0.125). Score 6 showed negative spectral features which could be assigned to proteins and amino acids (peaks at 904, 942 and 1003 cm^-1^) [37-39], higher (negative) for the Control group, and positive spectral features which may be assigned to sulfur compounds (C–S stretching – sulfides?) (703 cm^-1^) [39, 47], remnant amines/nucleic acids (1256 and 1435 cm^-1^) [38] and remnant carotenoids (1007, 1159 and 1522 cm^-1^) [38], higher for COVID-19 group, as seen by the PC 6 (statistically significant difference, *t*-test, *p* < 0.05). Therefore, the Control showed higher amount of amino acids and the COVID-19 group showed higher amounts of lipids, sulfur and carotenoids.

A recent study by Thomas et al. (2020) [48] showed a dysregulation of nitrogen metabolism in COVID-19 patients, with decreased circulating levels of most amino acids, except for tryptophan, and increased markers of oxidant stress (e.g. methionine sulfoxide, cystine), proteolysis, and kidney dysfunction (e.g. creatine, creatinine, polyamines). Increased circulating levels of glucose and free fatty acids were also observed; metabolite levels of these compounds correlated with clinical laboratory markers of inflammation and disease severity (i.e. IL-6 and C-reactive protein) and renal function (i.e. blood urea nitrogen) [48]. Higher levels of sulfides may be associated with the antiviral and anti-inflammatory action of this gas [49].

Phosphocholine has been found to be upregulated in positive COVID-19 patients, probably due to activated macrophage-mediated immunity, causing increasing in arachidonic acid, which is responsible for the increase in nitric oxide that macrophages use as a cytotoxic metabolite in order to destroy the microorganism in addition there is an increase in C reactive protein that binds to phosphocholine, activating the complement system and recruiting more phagocytes (inflammatory process) [41, 42, 44, 50, 51].

In a recent study of the proteomic and metabolomic characterization of serum from COVID-19 patients versus controls, Shen et al. (2020) [44] reported that more than 100 proteins were differentially expressed in COVID-19 positive patients compared to non-positive, where 50 of these proteins belonging to three pathways: activation of the complement system, macrophage function, and platelet degranulation. A number of metabolites (204) were correlated with disease severity and the most significantly changed metabolites were also involved in the same three biological processes. Therefore, the lower level of proteins and amino acids, and higher levels of lipids (including carotenoids), tryptophan, amines/amides and sulfur compounds seen in the Raman features are well aligned with the changes in the metabolism of positive patients.

Metabolites involved in arginine metabolism were also found to increase in COVID-19 patients [42, 45]. Therefore it may be expected an increase in the Raman features associated to compounds such as arginine, glutamine, glutamate and urea. The binding of SARS-CoV-2 to alveolar macrophages resulting in release of cytokines (interleukins) by macrophages [41]. These cytokines may appear as protein features, but these features were not seen in the positive samples probably due to low concentration.

Previous studies showed that Raman spectroscopy could be used to detect and quantify anti-Toxoplasma gondii IgG antibodies in sera of cats [52] and to detect dengue antibody IgM antibodies in human sera [53]. The Raman spectra were also used to detect hepatitis (B and C) virus infection in sera [54, 55] as well as dengue virus infection also in sera [56]. These studies of virus infections showed that hepatitis B presented difference in the metabolism of proteins, cholesterol, amino acids and nucleic acids due to liver infection [54] and hepatitis C presented difference related to sera components, but without specific biochemistry’s disease assignment [55]; the peaks at 1153 and 1510 cm^-1^ may have be erroneously assigned to carbohydrates and nucleic acids instead of carotenoids [54] and cytosine [55] in the control group. For the dengue virus patients, carotenoid peaks (1156 and 1516 cm^-1^) were decreased while peaks associated to immunoglobulins, adenosine diphosphate and hemoglobin were increased in positive patients [56]. Therefore different virus infection shows particular change in the biochemistry of the sera and consequently Raman features.

### 3.3 Discriminant analysis and classification

From the Raman spectra of sera and the PCs of both groups, classification models have been developed using discriminant analysis. The models were developed using the Chemoface application (www.ufla.br/chemoface) [32] using the leave-one-out cross-validation. Each spectrum was considered a sample in the discriminant models.

The models employed linear discriminant analysis (LDA) applied to selected PCs (the ones with significant difference between Control and COVID-19), and the PCA discriminant analysis (PCA-DA) and the PLS discriminant analysis (PLS-DA) applied to the entire spectrum, where the number of loading vectors (for the PCA-DA) and latent variables (for the PLS-DA) to be modeled were defined according to the higher discrimination accuracy with lower number of vectors or variables. Table 1 presents the results of the classification for each model in terms of sensitivity, specificity and accuracy. Better classification was achieved for the LDA using 4 PCs (PC2, PC4, PC5 and PC6), with 87% sensitivity and 100% specificity. Despite not significant (Figure 2), the PC 5 (urea) was essential for the classification obtained by LDA; without it, the correct classification lowered to 88% (data not shown).

**Table 1.** Sensitivity, specificity and percentage of correct classification (accuracy), for the classification of COVID-19 versus Control sera using the discriminations: LDA, PCA-DA and PLS-DA.

|  | LDA | PCA-DA | PLS-DA |
| --- | --- | --- | --- |
| No. of variables | 4 loadings* | 5 loadings | 5 latents |
| Sensitivity (%) | 87 | 77 | 80 |
| Specificity (%) | 100 | 100 | 100 |
| Accuracy (%) | 93 | 88 | 90 |

\* Loadings 2, 4, 5 and 6 (PC 2, PC 4, PC 5 and PC 6).

### 3.4 Future perspectives

It is extremely important that new technologies are used in the diagnosis of viral diseases, in order to make the diagnosis faster and more accurate, so that patients receive adequate and early treatment and minimize complications resulting from the disease. It is aligned with the need to develop a technique that has a lower cost per procedure for adoption as a screening technique in public health networks. The Raman technique can become a fast, accurate and less expensive methodology for the diagnosis of viral infection, as well as a possible use in the differential diagnosis between COVID-19 and seasonal flu (A/H1N1), as well as associating spectral changes with the degree of infection with COVID-19 (viral load).

Studies by other research groups are needed to confirm the spectral findings and to better correlate the spectral signatures of lipids, nitrogen compounds (amines/amides and urea) and nucleic acids found in COVID-19 group with the actual difference in the composition of the sera from the groups due to the virus. Also, it is important to increase the number of positive subjects versus controls in order to check the reproducibility of the spectra from both control and COVID-19 aiming the development of a technique for primary diagnosis as well as assessing the possibility of using the Raman technique for testing asymptomatic subjects on a large scale and assessing the presence of virus antibodies in serum, meeting the need for public health systems for the diagnosis of COVID-19 given the great need for identification and isolation of positive patients. The monitoring of positive patients and in resolution (cure) can also benefit from the advantages of the technique due to speed and lower cost.

The Raman spectral features reported for COVID-19 may be similar to other viral infections affecting the respiratory system such as seasonal influenza (A/H1N1, [57] and A/H3N2 [58]) employing surface enhanced Raman spectroscopy (SERS); therefore studies are needed to identify the particular features associated only to COVID-19 and to promote a differential diagnosis from infections by the seasonal flu.

## 4. Conclusion

The Raman spectroscopy applied to diagnose COVID-19 in human serum showed biochemical alterations related to the presence of the SARS-Cov-2 such as increase in lipids, nitrogen compounds (urea and amines/amides) and nucleic acids, and decrease in proteins and amino acids (tryptophan). The models based on discriminant analysis applied to the principal component loadings (PC 2, PC 4, PC 5 and PC 6) could classify spectra with 87% sensitivity, 100% specificity and 93% accuracy, demonstrating the possibilities of a rapid, label-free and costless technique for diagnosing COVID-19 infection.

## Data Availability

Data will be available upon request.

## Acknowledgements

A. C. C. Goulart thanks to Universidade Anhembi Morumbi for the master fellowship. L. Silveira Jr. acknowledges FAPESP (São Paulo Research Foundation) for granting the Raman spectrometer (Process 2009/01788-5) and CNPq (National Council for Scientific and Technological Development) for the productivity fellowship (Process No. 306344/2017-3).

